## Supplementary Information for "Impact of vaccinations, boosters and lockdowns on COVID-19 waves in French Polynesia"

#### 1 Supplementary methods

##### 1.1 Sensitivity analysis

In addition to the analysis with central parameter values presented in the main text, we refit the model and resimulate the counterfactual scenarios for pessimistic and optimistic assumptions about the key uncertain parameter values as a sensitivity analysis. Specifically, we vary the parameters shown in Table S1 across the central, pessimistic and optimistic scenarios.

Table S1: Parameter values used in sensitivity analysis

| Parameter assumptions | Booster waning rate, $\zeta_{i5}$ | Mean duration natural immunity, $1/\gamma_R$ | Cross-immunity to Delta from wild-type, $\eta_{Delta}$ | Cross-immunity to Omicron BA.1/BA.2 from wild-type/Delta, $\eta_{Omicron}$ | Vaccine effectiveness |
|---|---|---|---|---|---|
| Central | 0.0025 day$^{-1}$ [1] | 6 years [2–4] | 0.95 [4] | 0.55 [3] | central (Table S2) |
| Pessimistic | 0.0025 day$^{-1}$ [1] | 3 years [4, 5] | 0.75 [4] | 0.25 [6] | pessimistic (Table S2) |
| Optimistic | 0.00057 day$^{-1}$ [3] | 6 years [2–4] | 1 [4] | 0.55 [3] | optimistic (Table S2) |

Table S2: Vaccine effectiveness values used in sensitivity analysis

| Outcome | Dose | Vaccine effectiveness (%) | | | | | | | | | |
|---|---|---|---|---|---|---|---|---|---|---|---|
| | | Symbol | Wild-type/Alpha | | | Delta | | | Omicron BA.1/BA.2 | | |
| | | | Central* | Pessimistic† | Optimistic† | Central* | Pessimistic† | Optimistic† | Central* | Pessimistic‡ | Optimistic‡ |
| Infection | 1 | $e^{inf}$ | 70 | 63 | 77 | 62 | 55.8 | 68.2 | 34.2 | 34.2 | 34.2 |
| | 2 | | 85 | 76.5 | 93.5 | 80 | 72 | 88 | 44.1 | 44.1 | 44.1 |
| | 2 + waned | | 48 | 43.2 | 52.8 | 45 | 40.5 | 49.5 | 24.8 | 24.8 | 24.8 |
| | 2 + booster | | 95 | 85.5 | 99 | 91.4 | 82.26 | 99 | 65.9 | 45 | 65.9 |
| Symptoms | 1 | $e^{sympt}$ | 70 | 63 | 77 | 62 | 55.8 | 68.2 | 34.2 | 34.2 | 34.2 |
| | 2 | | 90 | 81 | 99 | 81 | 72.9 | 89.1 | 46.9 | 46.9 | 46.9 |
| | 2 + waned | | 51 | 45.9 | 56.1 | 61 | 54.9 | 67.1 | 46.7 | 46.7 | 46.7 |
| | 2 + booster | | 95 | 85.5 | 99 | 91.9 | 82.71 | 99 | 67.6 | 65 | 74 |
| Hospitalisation | 1 | $e^{SD}$ | 85 | 76.5 | 93.5 | 92 | 82.8 | 99 | 76.7 | 76.7 | 76.7 |
| | 2 | | 95 | 85.5 | 99 | 96 | 86.4 | 99 | 83.7 | 83.7 | 83.7 |
| | 2 + waned | | 65 | 58.5 | 71.5 | 84.2 | 75.78 | 92.62 | 67.6 | 67.6 | 67.6 |
| | 2 + booster | | 99 | 89.1 | 99 | 99 | 89.1 | 99 | 93.3 | 85 | 95 |
| Death | 1 | $e^{death}$ | 85 | 76.5 | 93.5 | 92 | 82.8 | 99 | 76.7 | 76.7 | 76.7 |
| | 2 | | 95 | 85.5 | 99 | 96 | 86.4 | 99 | 83.7 | 83.7 | 83.7 |
| | 2 + waned | | 66 | 59.4 | 72.6 | 84.2 | 75.78 | 92.62 | 67.6 | 67.6 | 67.6 |
| | 2 + booster | | 99 | 89.1 | 99 | 99 | 89.1 | 99 | 93.3 | 85 | 95 |
| Infectiousness if infected | 1 | $e^{ins}$ | 47 | 42.3 | 51.7 | 24 | 21.6 | 26.4 | 24 | 24 | 24 |
| | 2 | | 47 | 42.3 | 51.7 | 37 | 33.3 | 40.7 | 37 | 37 | 37 |
| | 2 + waned | | 30 | 27 | 33 | 24 | 21.6 | 26.4 | 24 | 24 | 24 |
| | 2 + booster | | 37 | 33.3 | 40.7 | 37 | 33.3 | 40.7 | 37 | 30 | 50 |

Sources:
*Barnard et al. [3]
†Assumed ±10%
‡Informed by Table 4a in [7]

### 2 Supplementary results

#### 2.1 MCMC output

The trace plots for the fitted parameters in Figure S1 show successful convergence of the MCMC, as the chains, which were started from different points in the 17-dimensional parameter space, all converge to the same values. Similarly, the maximum Gelman-Rubin diagnostic across all the parameters for the combined chains (4000 iterations) of 1.02 (which is well below the recommended threshold of 1.1) and the minimum effective sample size of 518 indicate that the chains have converged. The histograms of the posterior distributions for the parameters in Figure S2 show that they are all identifiable from the data. There is some positive correlation between the beta parameters for nearby time periods (Figure S3), which is not surprising given the piecewise linear form of the transmission rate (Equation (1) in the main text). The introduction dates for the wild-type virus and for the Delta variant ($t_0$ and $t_{Delta}$) are strongly positively correlated with the transmission parameters for the respective time periods ($\beta_1$ and $\beta_4$), which is to be expected given that the a higher transmission rate can compensate for a later introduction date. The maximum probability (over all age groups) of severe disease requiring hospitalisation given symptomatic infection ($p_{H\,max}$) is strongly negatively correlated with the relative risk of hospitalisation for the Delta variant over the wild-type virus ($\pi_{H\,Delta/Wildtype}$). This is due to the fact that nearly 60% of the hospitalisations occurred in the Delta wave, so a higher risk of hospitalisation given symptomatic infection can be counterbalanced by a lower relative risk of hospitalisation for Delta vs wild-type.

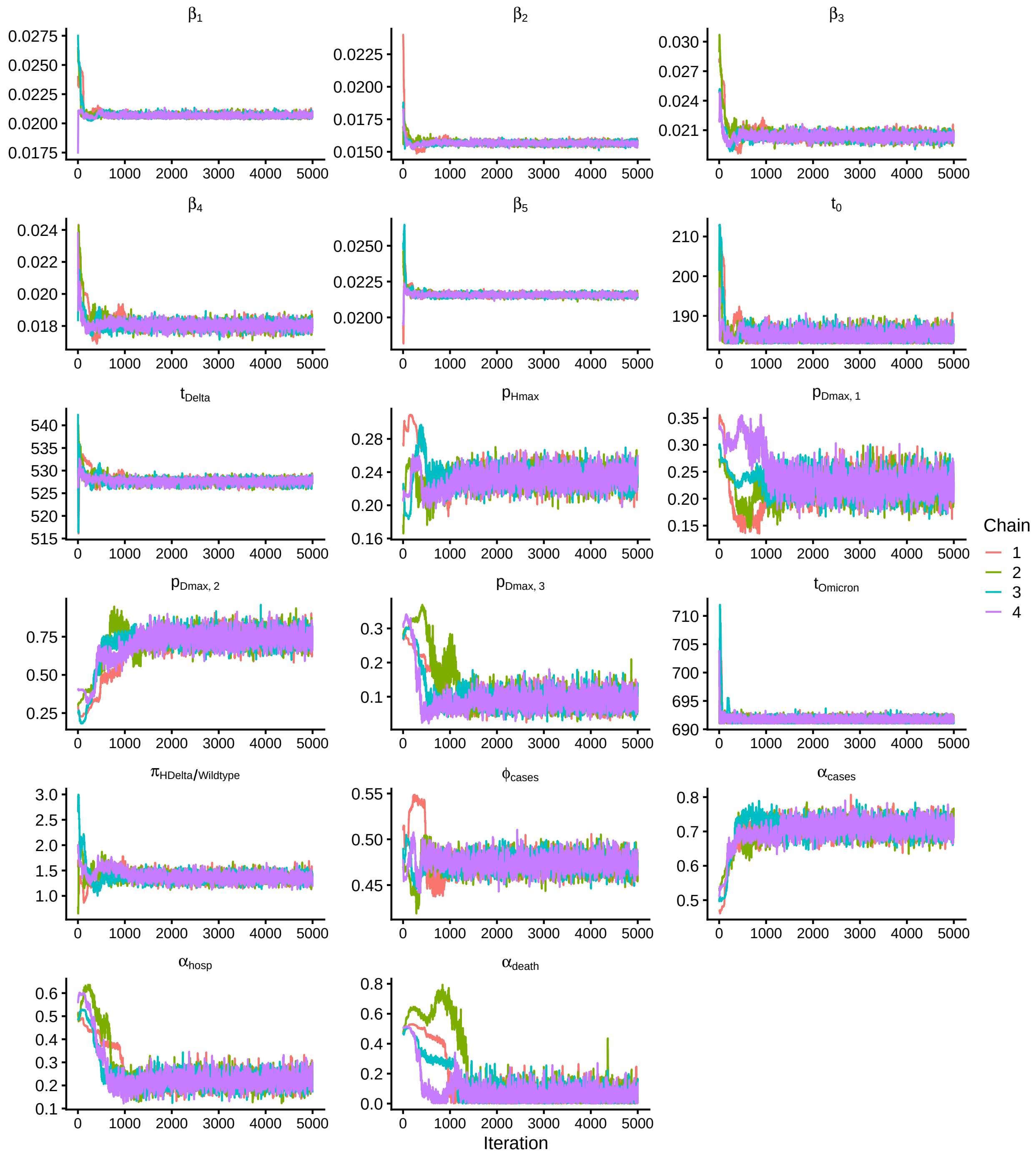


Figure S1: Trace plots for fitted parameters from MCMC. Plots show samples from full chain (50,000 iterations), including the burn-in, thinned by a factor of 10.

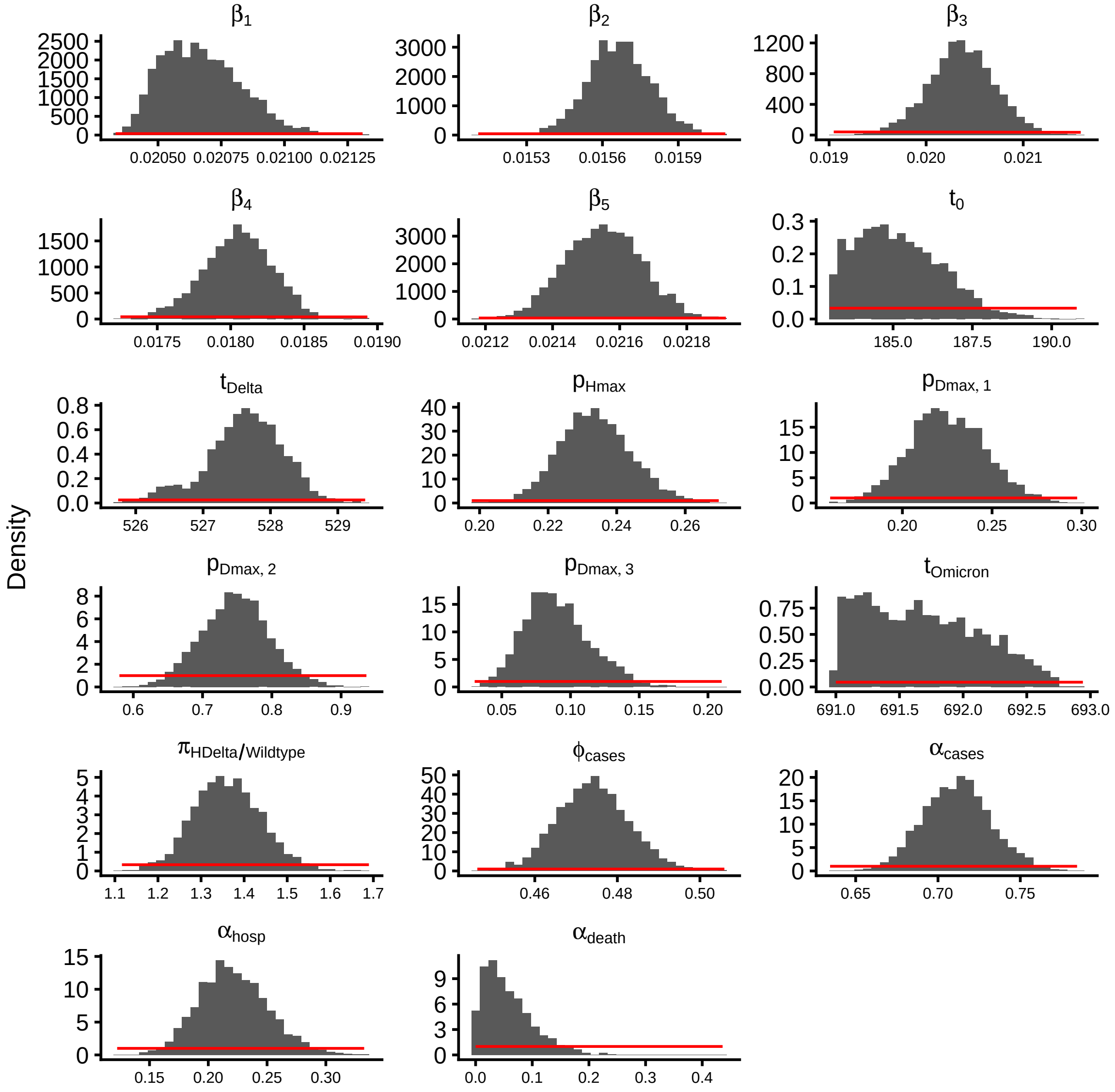


Figure S2: Prior (red lines) and posterior (histograms) distributions of fitted parameters

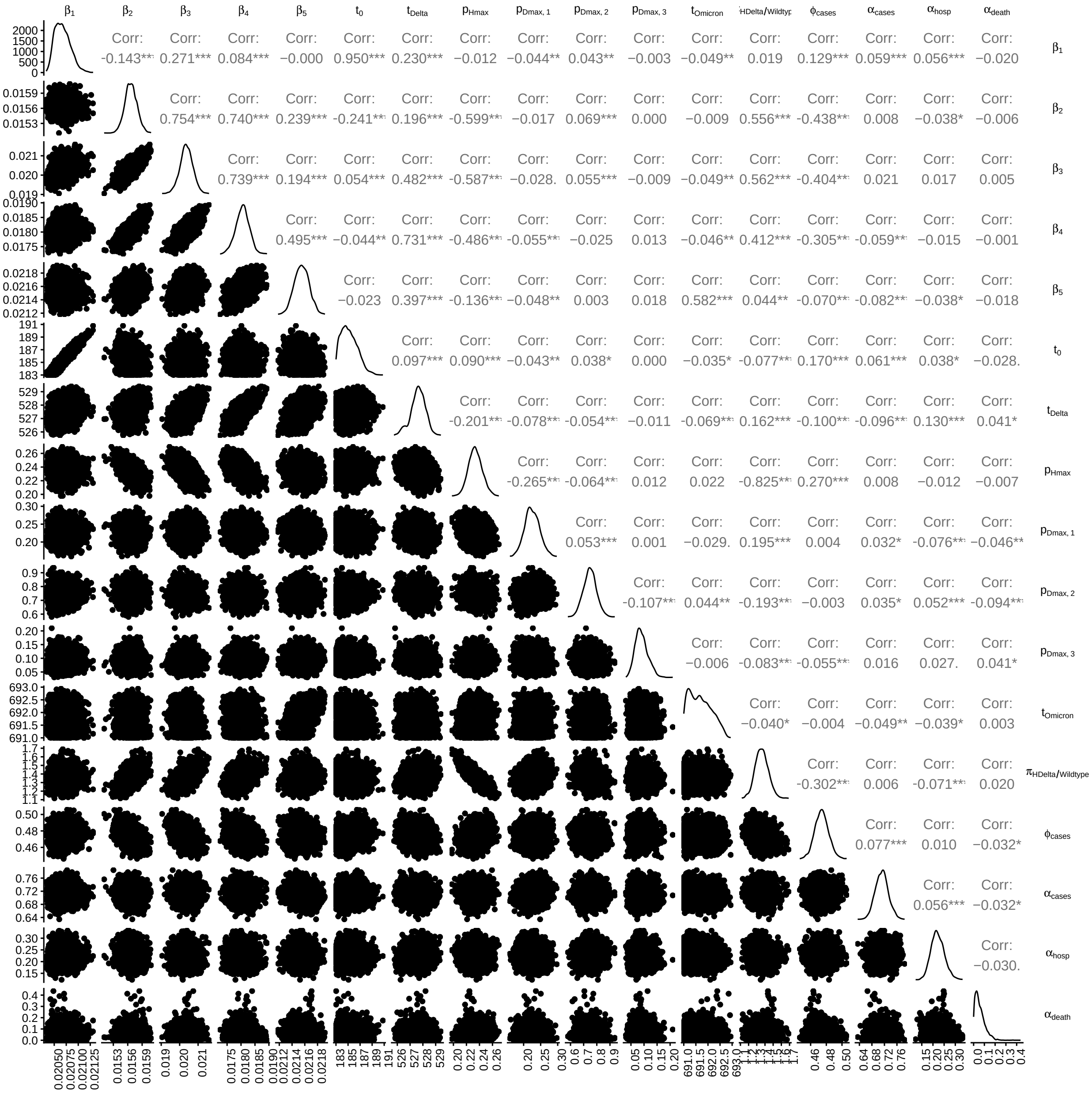

Figure S3: Posterior distributions and pairwise correlations of the fitted parameters. Parameter posterior distributions are shown on the diagonal, pairwise scatter plots in the lower triangle, and pairwise correlation coefficients in the upper triangle.

#### 2.2 Age-stratified model fit

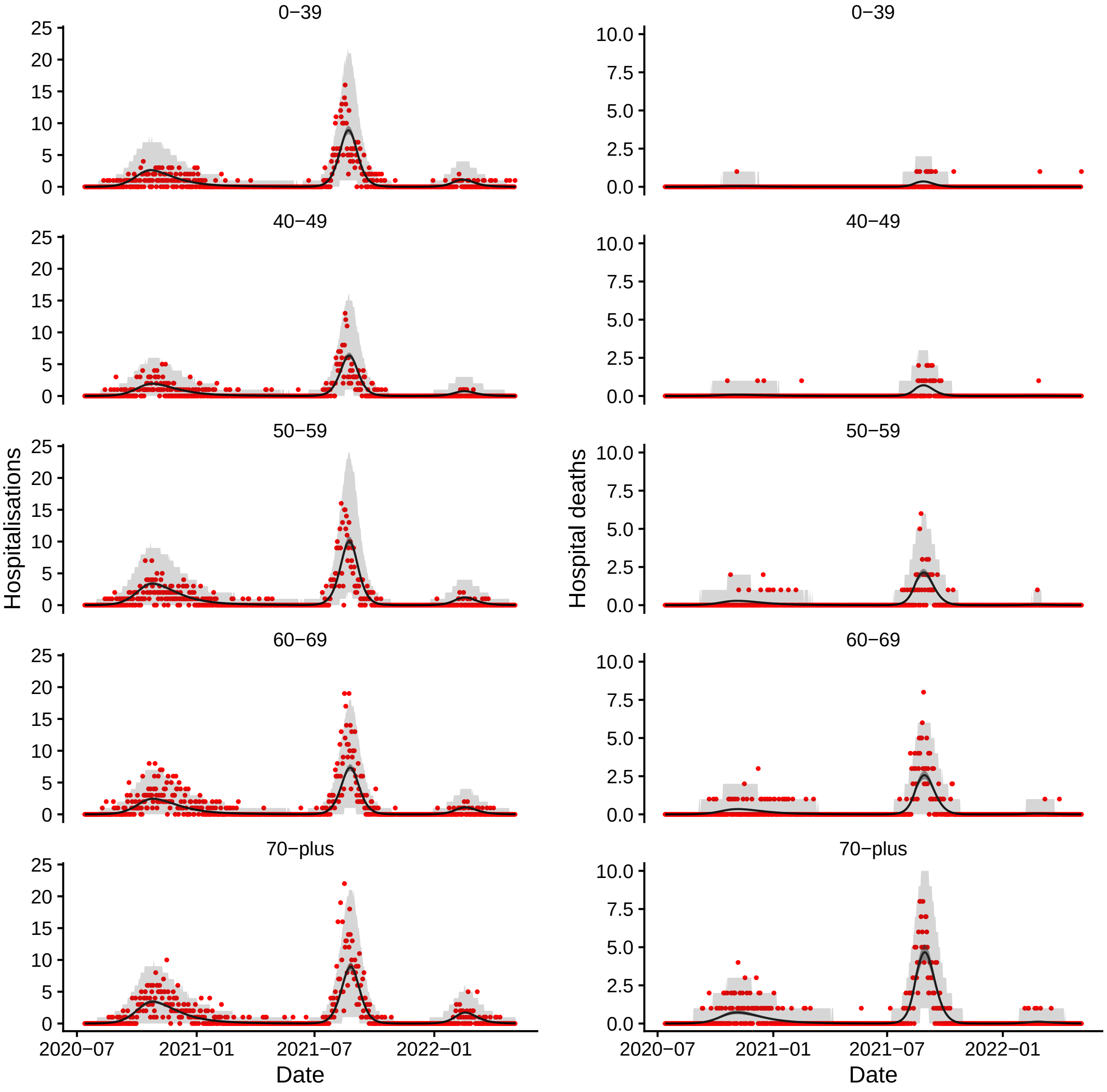


Figure S4: Fit of the model to age-stratified hospitalisation (left) and hospital death (right) data for French Polynesia from July 2020 to May 2022. Red points show data, black line and grey shaded area show median and 95% credible intervals of simulations of the fitted model, i.e. the uncertainty in the expected number of hospitalisations and deaths in the model. Light grey shaded area shows 95% posterior predictive interval of the model, i.e. the uncertainty in the hositalisations and deaths from the model also accounting for uncertainty in the observation process. Note different scales on vertical axes.

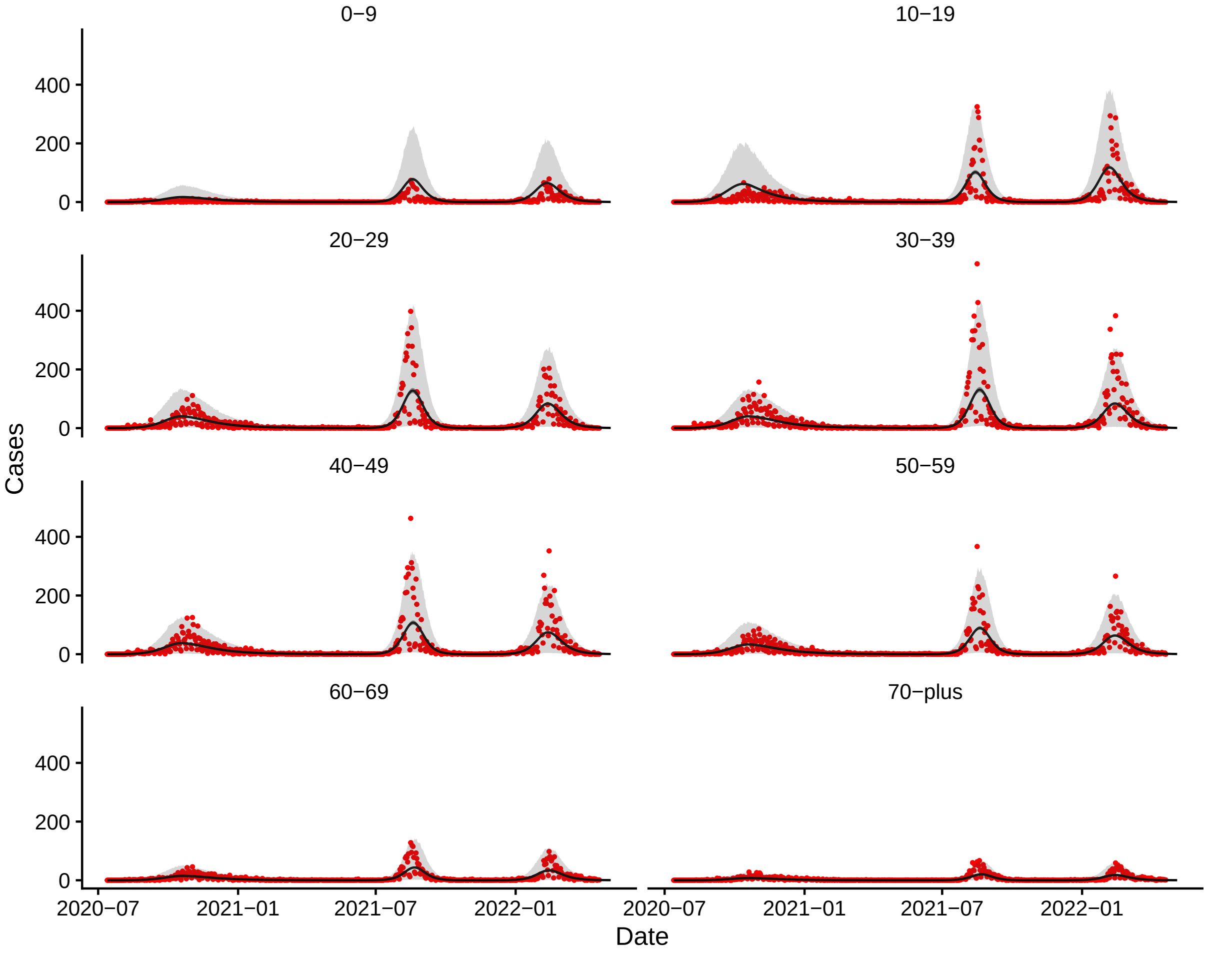


Figure S5: Fit of the model to age-stratified confirmed case data for French Polynesia from July 2020 to May 2022. Red points show data, black line and grey shaded area show median and 95% CI of simulations of the fitted model, i.e. the uncertainty in the expected number of cases in the model. Light grey shaded area shows 95% posterior predictive interval of the model, i.e. the uncertainty in the cases from the model also accounting for uncertainty in the observation process. Note that there is a strong day-of-the-week effect in the reporting.

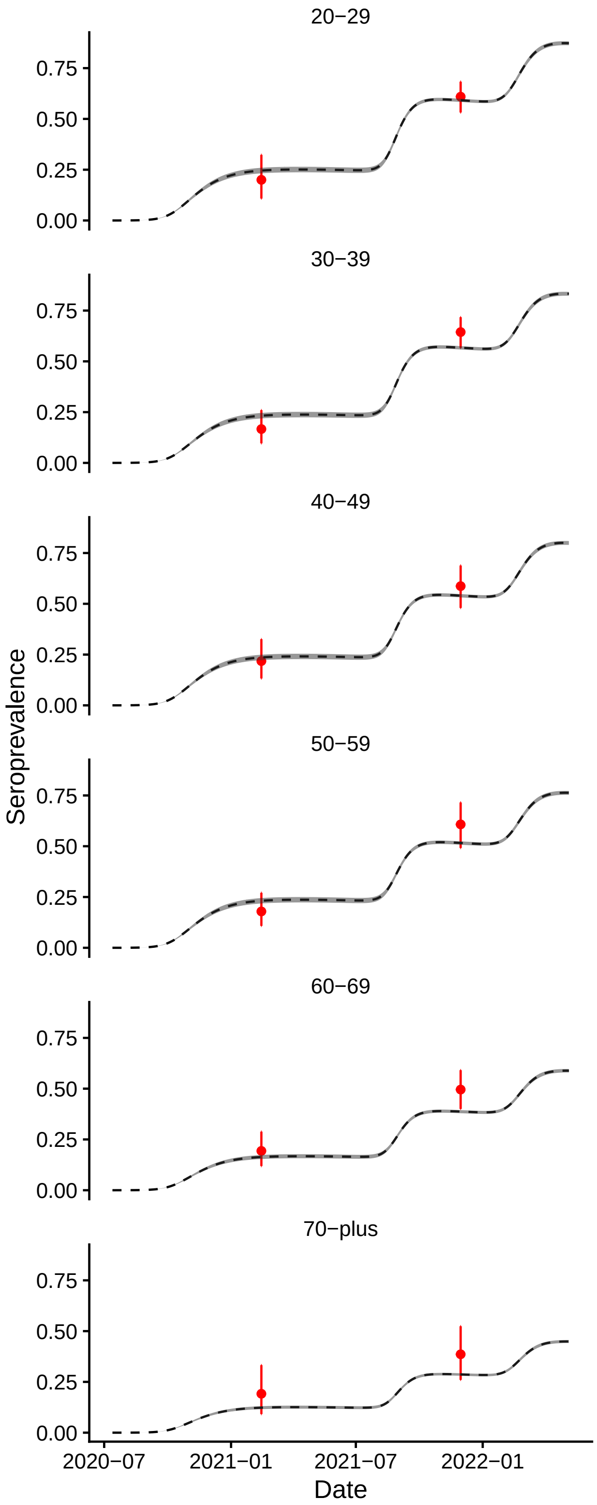


Figure S6: Fit of the model to age-stratified seroprevalence data from the two seroprevalence surveys conducted on Tahiti and Moorea by the Cellule Epi-surveillance COVID and the Health Department of French Polynesia in Feburary 2021, and on Tahiti by Institut Louis Malardé in November-December 2021. Red dots and bars show seroprevalence estimates and exact binomial 95% confidence intervals for the two sero-surveys, and black dashed line and grey shaded region show median and 95% CI of seroprevalence from simulations of the model.

Table S3: Median (95% CI) estimated total numbers of symptomatic cases, hospitalisations and hospital deaths in different COVID-19 waves in French Polynesia up to May 2022 for different counterfactual scenarios of lockdowns, vaccination and booster rollout.

| Counterfactual | Cases (thousands) | | | | Hospitalisations | | | | Hospital deaths | | | |
|---|---|---|---|---|---|---|---|---|---|---|---|---|
| | Wave 1 | Wave 2 | Wave 3 | Total | Wave 1 | Wave 2 | Wave 3 | Total | Wave 1 | Wave 2 | Wave 3 | Total |
| No change | 43.1 (40.8 – 45.4) | 52.5 (51 – 53.9) | 49.4 (48.7 – 50.2) | 145 (143 – 147) | 1170 (1090 – 1240) | 1500 (1420 – 1600) | 267 (252 – 284) | 2940 (2810 – 3080) | 132 (110 – 154) | 408 (364 – 448) | 10.7 (6.22 – 16.6) | 549 (499 – 600) |
| No lockdowns | 70.8 (68.9 – 72.8) | 22.3 (19.5 – 24) | 54 (52.9 – 55.2) | 147 (145 – 148) | 2030 (1860 – 2220) | 568 (478 – 644) | 301 (283 – 320) | 2890 (2730 – 3070) | 237 (196 – 282) | 106 (84.9 – 128) | 12.4 (7.23 – 19.7) | 356 (312 – 402) |
| No vaccination | 43.3 (41 – 45.6) | 81.5 (79.9 – 83.2) | 97.7 (97.1 – 98.4) | 223 (221 – 224) | 1170 (1100 – 1250) | 3730 (3530 – 3960) | 919 (868 – 976) | 5830 (5560 – 6140) | 133 (111 – 155) | 1370 (1230 – 1520) | 32.6 (18.9 – 50.3) | 1540 (1390 – 1680) |
| No boosters | 43.1 (40.8 – 45.4) | 52.5 (51.1 – 53.9) | 56.2 (55.5 – 57) | 152 (150 – 154) | 1170 (1090 – 1240) | 1500 (1420 – 1600) | 324 (305 – 344) | 2990 (2870 – 3140) | 132 (110 – 154) | 408 (364 – 448) | 13.2 (7.66 – 20.4) | 552 (502 – 602) |

Wave 1 = 13th July 2020 to 10th June 2021
Wave 2 = 11th June 2021 to 20th November 2021
Wave 3 = 21st November 2021 to 6th May 2022.

#### 2.3 Impact of alternative lockdown timings

We estimate the counterfactual impact that starting the lockdowns during the first two COVID-19 waves earlier or later would have had on the numbers of symptomatic cases, hospitalisations and hospital deaths in each wave and overall by simulating different combinations of the dates of the estimated changes in transmission due to the lockdowns in the first two waves (Figure S10). For each combination of lockdown dates, we run 500 simulations with parameter values drawn from the posterior distribution of the parameters from the model fitting and compare the numbers of symptomatic cases, hospitalisations and hospital deaths to those in simulations with the same parameter values with the actual lockdown dates. This gives the results shown in Table S4 and Figure S7.

The differences in the overall numbers of symptomatic cases, hospitalisations and hospital deaths for the earlier and later lockdown timings are all relatively small, ranging from 500 (95% CI 500–600) fewer cases for starting the first lockdown 2 weeks earlier, 35 (95% CI 17–54) fewer hospitalisations for starting both lockdowns 2 weeks earlier and 42 (95% CI 32–55) fewer deaths for starting the 1st lockdown 2 weeks later up to 200 (95% CI 100–400) more cases for starting both lockdowns 2 weeks later, and 18 (95% CI 6–31) more hospitalisations and 21 (95% CI 15–28) more deaths for starting the second lockdown 2 weeks later (Table S3). This is because starting the first lockdown either 2 weeks earlier or later would have increased the number of cases in the first wave — by 700 (95% CI 200–1100) and 3800 (95% CI 3100–4600) respectively — and a greater number of cases in the first wave would have resulted in greater accumulation of population immunity prior to the second wave and therefore fewer cases, hospitalisations and deaths in the second wave. Starting the lockdown earlier would have flattened the first wave (reduced its peak, but prolonged its duration), while starting it later would have increased the peak but shortened the duration (Figure S7). The former change would have resulted in 1600 (95% CI 900-2100) fewer cases in the second wave, the latter 4500 (95% CI 3600–5600) fewer cases. This bigger reduction in the number of cases in the second wave leads to a greater reduction in overall numbers of deaths from delaying the first lockdown than starting it earlier (even though the latter leads to the biggest reduction in cases), since there are more deaths per case in the second wave due to the greater severity of the Delta variant. The decrease in the overall number of hospitalisations is similar but slightly smaller (30 vs 35) than for starting both lockdowns 2 weeks earlier just due to the balance of the increase in the size of the first wave and the decrease in the size of the second wave under each scenario. Initiating the second lockdown 2 weeks later would have slightly increased hospitalisations and deaths overall as it would have led to a higher number of cases in the second wave. Varying the lockdown timings in the first two waves would have had a limited impact on the size of the third wave, as the total number of infections across the first two waves would have remained similar (so the population-level immunity entering the third wave would have been similar) and the immune escape properties of the Omicron BA.1/BA.2 variants reduce the impact of the proportion previously infected on the size of the third wave.

Table S4: Median (95% CI) estimated total numbers of symptomatic cases, hospitalisations and hospital deaths in different COVID-19 waves in French Polynesia up to May 2022 for different counterfactual scenarios of the timings of the lockdowns in the 1st and 2nd waves under 'central' parameter assumptions (Table S1).

| Counterfactual | Cases (thousands) | | | | Hospitalisations | | | | Hospital deaths | | | |
|---|---|---|---|---|---|---|---|---|---|---|---|---|
| | Wave 1 | Wave 2 | Wave 3 | Total | Wave 1 | Wave 2 | Wave 3 | Total | Wave 1 | Wave 2 | Wave 3 | Total |
| No change in lockdown dates | 43.1 (40.8 – 45.4) | 52.5 (51 – 53.9) | 49.4 (48.7 – 50.2) | 145 (143 – 147) | 1170 (1090 – 1240) | 1500 (1420 – 1600) | 267 (252 – 284) | 2940 (2810 – 3080) | 132 (110 – 154) | 408 (364 – 448) | 10.7 (6.22 – 16.6) | 549 (499 – 600) |
| 1st lockdown 2 weeks earlier | 43.8 (41 – 46.3) | 51 (49.1 – 52.9) | 49.7 (49 – 50.5) | 145 (143 – 146) | 1180 (1110 – 1250) | 1450 (1370 – 1550) | 269 (253 – 286) | 2900 (2780 – 3050) | 133 (111 – 155) | 388 (347 – 428) | 10.9 (6.29 – 16.8) | 531 (483 – 582) |
| 1st lockdown 2 weeks later | 46.9 (45.1 – 48.6) | 47.9 (46.7 – 49.1) | 50.3 (49.5 – 51) | 145 (143 – 147) | 1280 (1200 – 1380) | 1360 (1280 – 1440) | 273 (257 – 290) | 2910 (2780 – 3050) | 146 (121 – 171) | 350 (313 – 388) | 11 (6.39 – 17.1) | 507 (462 – 556) |
| 2nd lockdown 2 weeks earlier | 43.5 (41.2 – 45.8) | 52.6 (50.9 – 54.3) | 49.1 (48.4 – 49.8) | 145 (143 – 147) | 1180 (1100 – 1250) | 1500 (1410 – 1590) | 265 (249 – 281) | 2940 (2810 – 3090) | 134 (111 – 156) | 398 (355 – 436) | 10.6 (6.16 – 16.4) | 540 (490 – 592) |
| 2nd lockdown 2 weeks later | 42.8 (40.5 – 45.1) | 52.9 (51.8 – 54.1) | 49.5 (48.6 – 50.3) | 145 (143 – 147) | 1160 (1080 – 1230) | 1530 (1450 – 1620) | 267 (252 – 284) | 2960 (2830 – 3100) | 131 (109 – 153) | 430 (384 – 473) | 10.7 (6.21 – 16.6) | 572 (519 – 624) |
| Both lockdowns 2 weeks earlier | 44.5 (41.7 – 47.1) | 50.6 (48.4 – 52.8) | 49.5 (48.8 – 50.3) | 145 (143 – 146) | 1200 (1120 – 1280) | 1430 (1340 – 1540) | 268 (252 – 285) | 2900 (2770 – 3050) | 136 (113 – 158) | 372 (330 – 411) | 10.8 (6.25 – 16.7) | 516 (469 – 569) |
| Both lockdowns 2 weeks later | 46.8 (45 – 48.5) | 48 (47 – 49.1) | 50.4 (49.6 – 51.2) | 145 (143 – 147) | 1280 (1200 – 1370) | 1370 (1290 – 1460) | 274 (258 – 291) | 2920 (2800 – 3070) | 146 (121 – 171) | 368 (328 – 407) | 11 (6.41 – 17.1) | 524 (477 – 575) |

Wave 1 = 13th July 2020 to 10th June 2021
Wave 2 = 11th June 2021 to 20th November 2021
Wave 3 = 21st November 2021 to 6th May 2022.

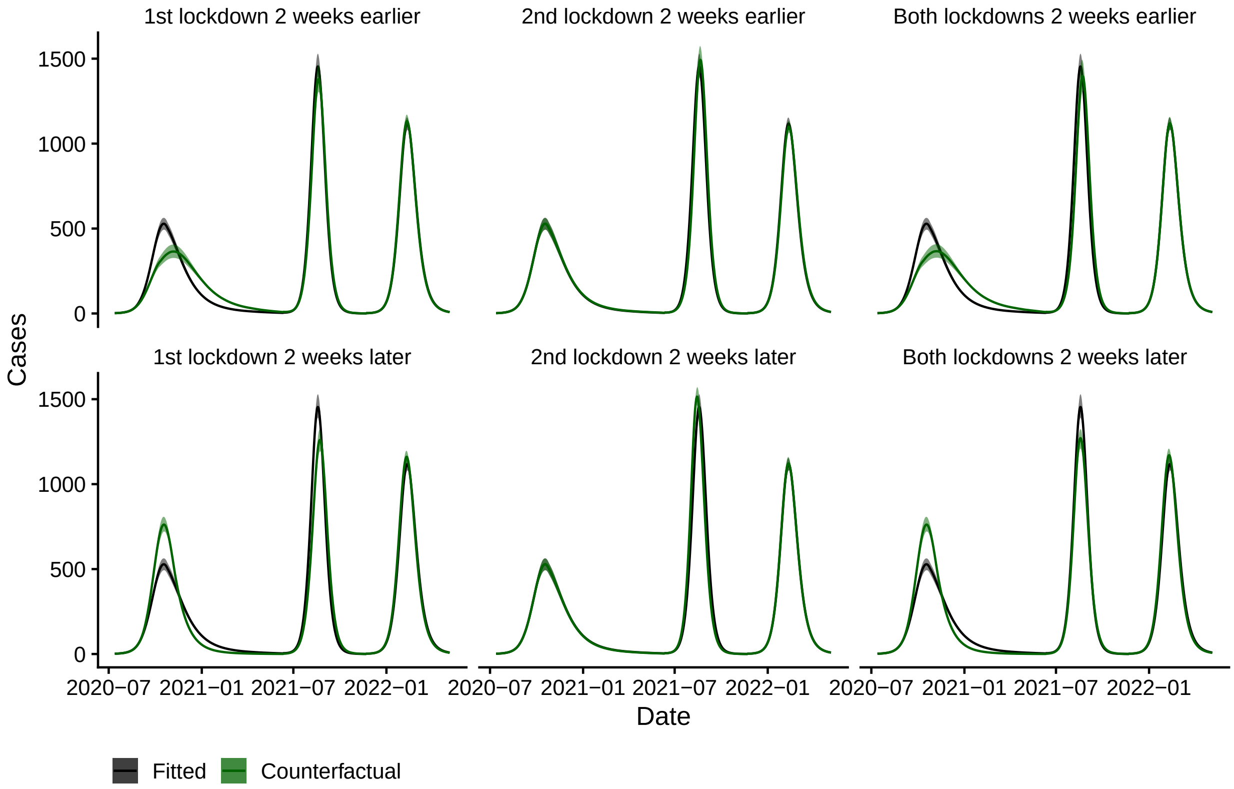


Figure S7: Impact of alternative lockdown scenarios on numbers of COVID-19 symptomatic cases under 'central' parameter assumptions.

#### 2.4 Sensitivity analysis

Figure S8 shows the impact of the different assumptions for uncertain parameter values on the estimated numbers of symptomatic cases, hospitalisations, and hospital deaths averted between July 2020 and May 2022 as a result of different interventions (lockdowns, all vaccination, boosters).

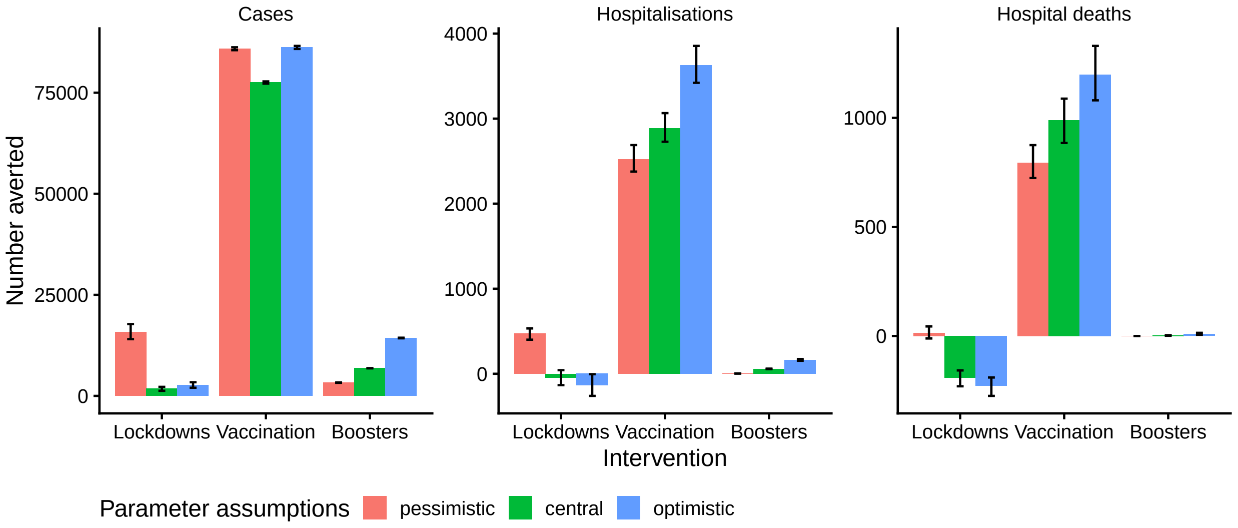


Figure S8: Estimated numbers of symptomatic cases, hospitalisations, and hospital deaths averted under different assumptions about uncertain parameters (see Table S1). Error bars show 95% CIs. Note different scales on the vertical axes.

Varying the parameter assumptions has a nonlinear effect on the estimated number of cases averted through lockdowns. Fewest cases are estimated to have been averted under the central assumptions (1800, 95% CI 1300–2200), then the optimistic assumptions (2700, 95% CI 2000–3400), and then the pessimistic assumptions (15,800, 95% CI 14,000–17,700). This is because of the impact of immunity from infection in previous waves on the size of the second and third waves. Under the central assumptions, the larger first wave without lockdown results in a smaller second wave than actually occurred, but a similar sized third wave (Figure S9, top left panel, green curve and black dashed curve). Under the optimistic assumptions, however, the higher assumed cross-immunity from wild-type infection and higher vaccine effectiveness lead to a very small second wave, which in turn leads to less immunity and a larger third wave (and more cases overall than under the central assumptions) (Figure S9, top left panel, blue curve). Under the pessimistic assumptions the lower cross-immunity from previous infection, faster waning of immunity, and lower vaccine effectiveness result in similar sized second and third waves to those that actually occurred (Figure S9, top left panel, red curve and black dashed curve), which together with the larger first wave gives the much larger estimate of cases averted. Under the pessimistic assumptions removing lockdowns increases hospitalisations and hospital deaths by 472 (95% CI 401–533) and 15 (95% CI -11–44) respectively, since the first wave is larger and the later waves are largely unchanged (Figure S9, top middle and top right panels, red curves and black dashed curves); while under the central and optimistic assumptions it reduces hospitalisations by 45 (95% CI -42–134) and 136 (95% CI 5–260), and deaths by 193 (95% CI 158–231) and 229 (95% CI 190–274) respectively since the larger first (wild-type) wave is counterbalanced by the smaller second (Delta) wave (Figure S9, top middle and top right panels, green and blue curves) and there are fewer hospitalisations and deaths per case for wild-type than Delta as it has lower severity.

As for the central parameter assumptions, for the pessimistic and optimistic parameter assumptions, vaccination is estimated to have caused by far the largest reduction in cases, hospitalisations and deaths out of the different interventions. The effect of the different parameter assumptions on the estimated number of cases averted through vaccination is non-linear. The number averted is smallest under the central assumptions (77,500, 95% CI 77,200–77,800), then under the pessimistic assumptions (85,900, 95% CI 85,500–86,200), and then under the optimistic assumptions (86,200, 95% CI 85,800–86,600), though the differences are relatively small. This is due to the combined nonlinear effects on infection incidence of the differences in the rate of waning of natural and booster-derived immunity, cross-immunity to infection with new variants, and transmission rate inferred from the observed data between the different parameter assumptions. The estimated numbers of hospitalisations and hospital deaths averted through vaccination range from 2520 (95% CI 2380–2690) and 794 (95% CI 724–875) under the pessimistic assumptions to 3630 (95% CI 3420–3860) and 1197 (95% CI 1080–1330) under the optimistic parameter assumptions, due to the increase in vaccine effectiveness, duration of immunity, and levels of cross-immunity across the assumptions.

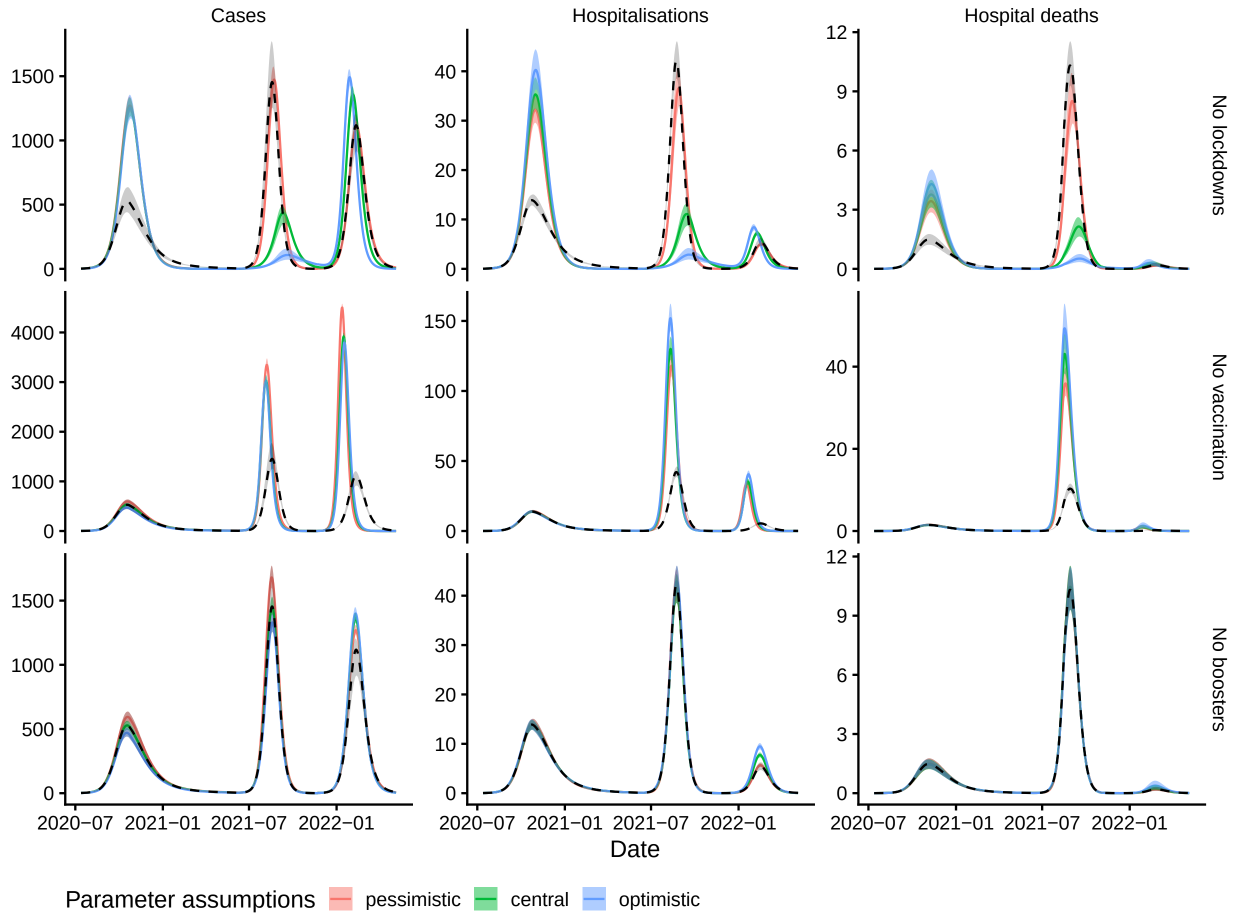


Figure S9: Numbers of cases, hospitalisations, and hospital deaths (columns) under different counterfactual scenarios (rows) for different assumptions about uncertain parameters (see Table S1). Solid lines and shaded areas show medians and 95% CI of 500 simulations of the model. Dashed black line and grey shaded area show median and 95% CI of simulations of the fitted model across the different parameter assumptions. Note different scales on the vertical axes.

Booster doses are estimated to have averted between 3300 (95% CI 3200–3300) and 14,300 (95% CI 14,200–14,400) cases, between 3 (95% CI 3–3) and 163 (95% CI 153–174) hospitalisations, and between 0 (95% CI 0–0) and 9 (95% CI 6–15) hospital deaths under the different parameter assumptions (going from pessimistic to optimistic) (Figure S8 and Figure S9 bottom row). The variation across the different parameter assumptions is relatively small – across all assumptions boosters are estimated to have contributed relatively little to reducing hospitalisations and deaths compared with first and second dose vaccination — and it is driven mainly by the different assumed booster waning rates, since these determine how quickly individuals lose all protection from boosters and return to being completely susceptible.

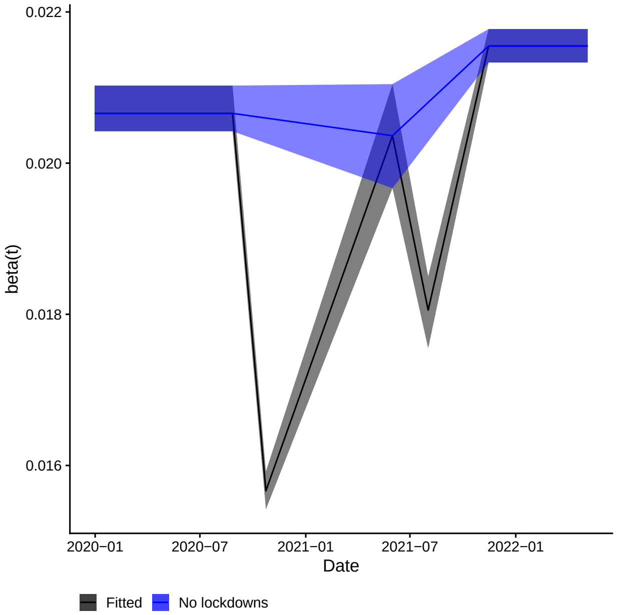


Figure S10: Estimated transmission rate over time, $\beta(t)$ under 'central' parameter assumptions. Black line shows median estimate, shaded region the 95% CI for the fitted model. Blue line and shaded region show the same for the "no lockdowns" counterfactual scenario.

### 3 Supplementary discussion

#### 3.1 Limitations

There are some factors that may mean we underestimate the impact of the vaccination programme. These include the fact that our estimates of cases, hospitalisations and deaths averted are based on reported hospital deaths, while data on all-cause mortality for 2015-2021 [8] and model-based estimates of excess mortality [9] suggest there may have been considerable under-reporting of COVID-19 deaths in the Delta wave. Although we fit the maximum probability of death given hospitalisation in each wave and thus account for the considerable variation in the risk of hospital death over time, we still underestimate the number of hospital deaths among individuals aged 60-69 years during the Delta wave (by approximately 20%). The increased risk of death in this age group in the Delta wave may be related to the main hospitals reaching capacity at its peak and it not being possible to provide the highest risk patients with the same level of care. We do not account for any such age-specific effect of hospital capacity in the model. Hospitals reaching capacity may also have led to increased death rates in the community (since some individuals with severe disease would not have been admitted). As the model does not account for this, we may have underestimated the transmission rate during this period and thus the counterfactual number of deaths without vaccination (as hospital capacity in the counterfactual scenario would have been reached more quickly, leading to a faster accumulation of deaths in the community).

Some of the large uncertainty in the impact of the booster programme (3300–14,300 symptomatic cases averted) stems from the structure of the model, in that we treat individuals whose booster protection wanes as returning to full susceptibility when they may in fact retain some protection against severe disease for a long period of time. However, we made this simplifying assumption due to the relatively limited data available on the rate at which booster protection wanes and to avoid increasing the complexity of the model further (e.g. with additional strata for waned booster protection), which would require more assumptions. Although the booster waning rate is based on data on the rate at which booster protection against Omicron BA.1 hospitalisation wanes [1], the estimate of booster impact at the lower end of the range may therefore still be too pessimistic.

Another potential source of underestimation of the impact of the booster programme is that we treat the Omicron BA.1 and BA.2 sublineages as the same, despite evidence that BA.2 is more transmissible than BA.1 [10–13] and can reinfect individuals previously infected with BA.1 [14], and may therefore cause higher infection rates in the absence of protection from boosters. To some extent the difference in transmissibility will be absorbed in the fitting of the transmission rate for the third wave, but the resulting averaged transmission rate will give a lower estimate for the number of cases, hospitalisations and deaths averted in the latter half of the third wave when BA.2 was dominant and booster coverage was higher. Despite the potential for reinfection with BA.2 following BA.1 infection, estimates of the rate of such reinfection are reasonably low (6–30% [14]), and there is evidence that infection with BA.1 elicits strong neutralising antibody responses against infection with BA.2 (Supplementary Figure 2 [15]). Vaccine effectiveness and severity estimates for BA.2 are also similar to those for BA.1 [16–20]. Therefore, given the lower severity of the Omicron variants, the absolute error in numbers of hospitalisations and deaths averted is likely to be small.

Finally, we have not fitted the model to the most recent waves in French Polynesia, caused mainly by the Omicron BA.5, BA.4, and BQ.1.1 variants, as doing so would require increasing the complexity of the model (or moving to a status-based approach to capturing the immune status of the population [21]) to account for repeat booster vaccinations and the increased transmissibility/immune escape of the BA.5 [22] and BQ.1.1 variants [23]. Hence we are not able to make projections of future incidence under different scenarios of increasing contact levels and introduction of new variants.
